# Development of patient-reported outcome for spinal and bulbar muscular atrophy

**DOI:** 10.64898/2026.07.02.26356783

**Authors:** Miho Nagae, Shinichiro Yamada, Daisuke Ito, Yoshiyuki Kishimoto, Shota Komori, Takahiro Kawase, Madoka Iida, Kondo Ayano, Munetaka Yamamoto, Abdullah Alqahtani, Narjis Kazmi, Christopher Grunseich, Masahisa Katsuno, Atsushi Hashizume

## Abstract

**Objectives:** To develop and validate a disease-specific patient-reported outcome (PRO) measure for spinal and bulbar muscular atrophy (SBMA).

**Methods:** A three-stage sequential design was adopted. Items were generated through qualitative interviews with patients with SBMA and expert review, refined using quantitative analyses, and evaluated for reliability and validity in independent cohorts from Japan and the United States.

**Results:** Interviews with 12 patients generated 234 candidate items, which were refined into a final 31-item SBMAPRO comprising five domains based on an online survey of 106 patients. Internal consistency across domains ranged from Cronbach’s alpha values of 0.651 to 0.901. In the Japanese cohort, test–retest reliability yielded intraclass correlation coefficients of 0.941 for physical function, 0.877 for mental health, and 0.858 for social function. Construct validity was examined through correlations with disease-specific functional measurements and the 36-Item Short Form Survey (SF-36). The SBMAPRO correlated with the SBMA Functional Rating Scale (*r* = −0.826, *p* <0.001) and with the SF-36 mental health (*r* = −0.693, *p* <0.001) and social functioning (*r* = −0.617, *p* <0.001) domains. In subscale analyses, the SBMAPRO social domain was associated with trunk–lower limb-related functional impairment (*r* = −0.587, *p* < 0.001). Similar patterns were observed in the American cohort.

**Conclusion:** The SBMAPRO demonstrated reliability and validity in Japanese and American cohorts. Associations between mental and social domains and trunk–lower limb dysfunction suggest that mobility impairment may contribute to psychological burden and restricted social participation in SBMA, indicating that this disease-specific PRO may complement clinician-rated measures.

## Introduction

Spinal and bulbar muscular atrophy (SBMA) is an inherited neuromuscular disease caused by expansion of a CAG trinucleotide repeat in the androgen receptor gene on the X chromosome. In 1991, La Spada et al. reported abnormal elongation of the CAG repeat sequence in exon 1 of the androgen receptor (*AR*) gene located on Xq11–q12 in patients with SBMA, a finding that initiated extensive investigations into the disease’s pathophysiology [1].

The estimated prevalence of SBMA is 1–2 per 100,000 individuals [2]. Clinical manifestations include muscle weakness affecting both proximal and distal limb muscles, fasciculations, muscle atrophy, tremors, sensory disturbances, and bulbar muscle involvement. Non-motor features such as gynecomastia, erectile dysfunction, testicular atrophy, and metabolic abnormalities are also commonly observed [3, 4, 5]. Neurological symptoms typically begin between 30 and 50 years of age [6, 7].

Although SBMA progresses relatively slowly, patients experience steady and cumulative functional decline over decades. Progressive impairment of limb and bulbar function leads to difficulties in mobility, speech, and swallowing, ultimately resulting in a substantial impact on activities of daily living (ADL) and overall quality of life (QoL). Severely affected individuals are at increased risk of aspiration pneumonia and ventilatory failure due to weakness of the bulbar and respiratory musculature [8, 9].

Despite a relatively preserved life expectancy, SBMA imposes a substantial and multifaceted disease burden. Functional decline negatively affects ADL and QoL. The reported median intervals from symptom onset are approximately 5 years to the need for handrail support, 6 years to the development of dysarthria, and 10 years to dysphagia [9]. To assess physical function in SBMA, disease-specific functional rating scales such as the SBMA Functional Rating Scale (SBMAFRS) [10] and the SBMA Functional Composite (SBMAFC) [11] have been developed and are used as quantitative indicators of overall disability and outcome measures in clinical research. However, these instruments primarily assess observable motor performance and do not fully capture the psychological, social, and subjective burdens experienced by patients.

Recently, value-based healthcare has increasingly emphasized patient-centered outcomes, including health-related quality of life (HR-QoL), in addition to traditional objective indicators such as survival or functional scores. Patient-reported outcomes (PROs) enable direct assessment of patients’ health experiences without physician interpretation and are advantageous in terms of time efficiency and cost-effectiveness. Regulatory authorities, such as the U.S. Food and Drug Administration and the European Medicines Agency, recommend incorporating PRO measures into clinical trials [12, 13].

Generic QoL instruments do not adequately capture SBMA-specific symptoms and may fail to detect subtle but meaningful changes perceived by patients, particularly in slowly progressive diseases such as SBMA. Because patients experience changes not only in clinician-assessed functional deterioration but also in various domains, including speech, swallowing, fatigue, mental health, and social participation, using an SBMA-specific PRO may allow for more accurate assessment of aspects of the disease that matter most to patients. To address this unmet need, we developed an SBMA-specific PRO measure, the SBMAPRO. This study describes the development of the SBMAPRO and evaluates its reliability and validity in Japanese and U.S. patient cohorts.

## Methods

### Study Design and Overview

The development and validation of the SBMAPRO followed a mixed-methods approach consisting of three sequential stages: item generation, item selection and instrument refinement, and psychometric evaluation. An overview of the development process is shown in Figure 1.

**Fig. 1.**
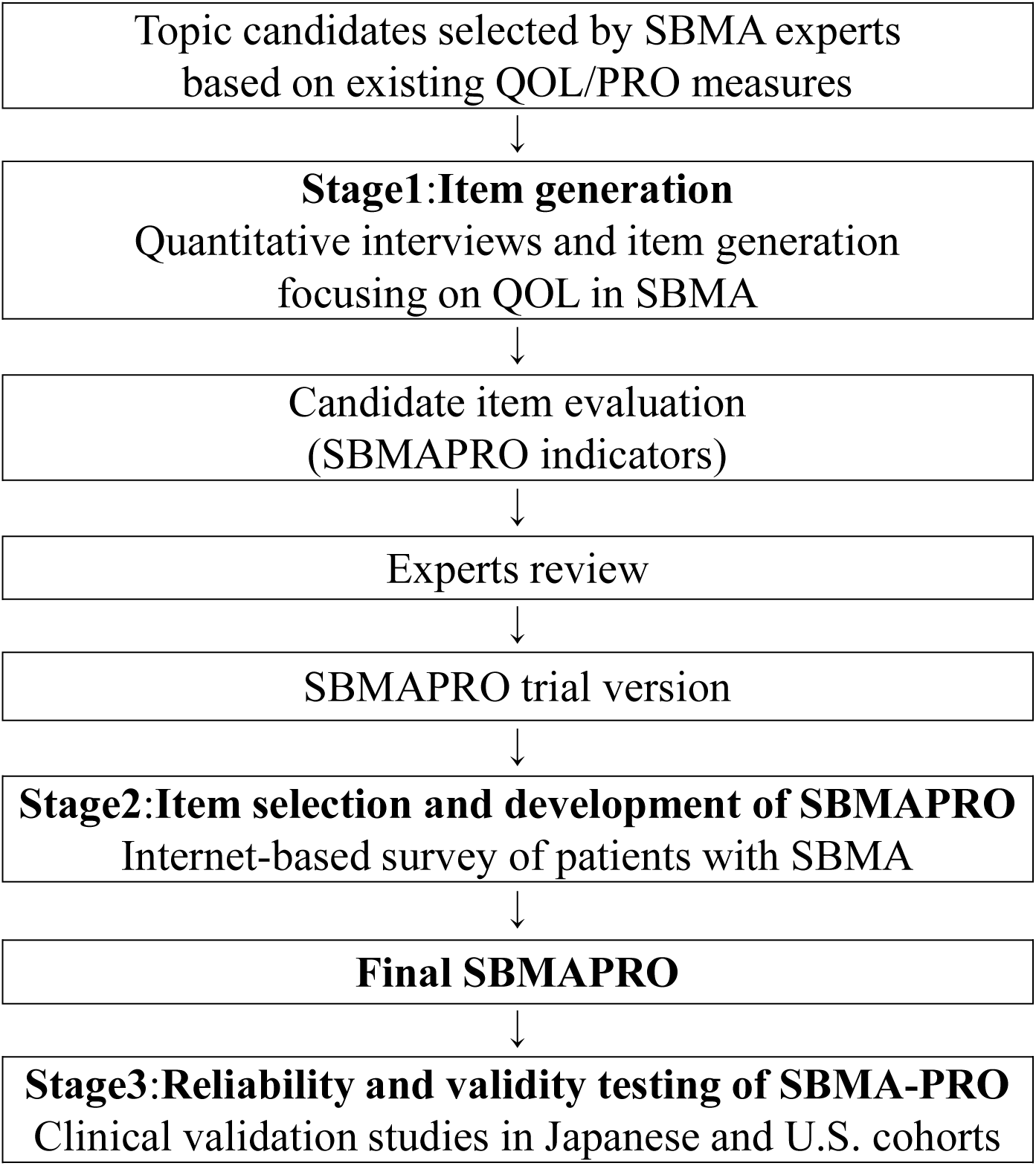
Overview of the SBMA-PRO development process. Schematic overview of the three-stage mixed-method approach used for the development and validation of the SBMA-PRO, including item generation, item selection and refinement, and psychometric evaluation

### Stage 1: Item Generation

#### Participants

Adults aged ≥20 years with a confirmed diagnosis of SBMA were eligible for participation. Participants were recruited from Nagoya University Hospital. Patients with severe complications (e.g., aspiration pneumonia, severe dysphagia, or respiratory insufficiency) that could substantially interfere with study participation were excluded [6, 8, 9]. The same inclusion and exclusion criteria were applied in Stage 2 and in the Japanese cohort of Stage 3. Procedures Semi-structured qualitative interviews were conducted with 12 patients with SBMA to comprehensively capture the impact of the disease on HR-QoL. Interviews addressed three predefined domains: (1) physical function, including limb function, speech and swallowing, sleep-related functions, excretory and sexual function, and fatigue; (2) mental health, including emotional changes, cognitive concerns, and anxiety about the future; and (3) social functioning, including changes in social roles and family relationships. Participants provided free-text descriptions of their experiences within each domain. Interview data were compiled into an initial item pool. Subsequently, SBMA experts reviewed all items to assess clinical relevance, clarity, and content validity and to remove redundant or ambiguous items.

### Stage 2: Item Selection and Development of the SBMAPRO

#### Participants

The target sample size was set at approximately 100 participants to ensure sufficient data for exploratory factor analysis of the SBMAPRO trial version. Procedures A cross-sectional, anonymous online survey was conducted to refine the draft items generated in Stage 1. The questionnaire was distributed via a secure online platform, and completion of the survey was considered to indicate informed consent. Statistical analysis The properties of the SBMAPRO draft were evaluated in a stepwise manner. Descriptive item and exploratory factor analyses were conducted to examine the underlying factor structure. Internal consistency of each domain was evaluated using Cronbach’s alpha coefficients. Responsiveness and interpretability were examined based on score distributions and the frequency of extreme responses. Statistical analyses were performed using IBM SPSS Statistics (Version 31.0.1.0).

### Stage 3: Reliability and Validity Testing of the SBMAPRO

#### Participants

In Japan, the study was designed to enroll 60 adults aged ≥20 years with a confirmed diagnosis of SBMA. In the United States, 25 adults with a confirmed diagnosis of SBMA were recruited from the Neurogenetics Branch of the National Institute of Neurological Disorders and Stroke, National Institutes of Health (Bethesda, MD, USA).

In the Japanese cohort, the target sample size was set at 60 participants based on an expected correlation coefficient of 0.80 and a confidence interval width of 0.20, allowing for potential dropouts [14]. In the United States, the study was conducted on a feasibility basis, and no predefined sample size calculation was performed.

#### Procedures

To assess test–retest reliability, Japanese participants completed the SBMAPRO at baseline and again after 1–2 weeks. This test-retest interval was set to minimize recall and practice effects while assuming stability of patients’ clinical status and to minimize the possibility of true change in the construct being measured, in line with established recommendations for PRO measures [15, 16].

To evaluate construct validity, participants in both the Japanese and U.S. cohorts completed the SBMAPRO, SBMAFRS, ALS Functional Rating Scale–Revised (ALSFRS-R), and the 36-Item Short Form Health Survey (SF-36) during outpatient visits.

#### Outcome measures

SBMA-specific functional impairment was assessed using the SBMAFRS, a disease-specific, clinician-rated scale developed to quantify motor and bulbar dysfunction in patients with SBMA [10]. Global functional status was evaluated using the ALSFRS-R, a widely used functional scale originally developed for amyotrophic lateral sclerosis and commonly applied across neuromuscular diseases [17]. HR-QoL was assessed using the SF-36 [18–21]. All outcome measurements were performed identically in both cohorts.

#### Statistical analysis

Test–retest reliability was assessed using intraclass correlation coefficients (ICCs), calculated using a two-way random-effects model with absolute agreement. Construct validity was evaluated by calculating Pearson’s correlation coefficients between the SBMAPRO scores and scores on the SBMAFRS, ALSFRS-R, and SF-36. Analyses were conducted separately for the Japanese and U.S. cohorts.

The strength of correlations was interpreted according to commonly used guidelines in medical research. Correlation coefficients of approximately 0.30–0.49 were considered to indicate moderate correlations, values of 0.50–0.69 to indicate moderate-to-strong correlations, and values of 0.70 or higher to indicate strong correlations, based on the criteria proposed by Altman [22]. Correlations between the mental and social domain scores of the SBMAPRO and SBMAFRS total and subscale scores were examined separately in the Japanese and U.S. cohorts.

Statistical analyses were performed using IBM SPSS Statistics (Version 31.0.1.0 (49)).

### Ethics

This study was approved by the Nagoya University Hospital Ethics Committee (approval number: 2022-0024). The study conducted in the United States was approved by the National Institutes of Health Institutional Review Board (protocol NCT04944940).

Written informed consent or electronic consent was obtained from all participants prior to enrollment.

## Results

### Stage 1: Item Generation

#### Participants

In Stage 1, 12 Japanese patients diagnosed with SBMA participated in a qualitative study based on semi-structured interviews. Participants ranged in age from 46 to 73 years. Development of the SBMAPRO Trial Version A total of 234 candidate items were generated, including 145 related to physical function, 54 to mental health, and 35 to social functioning. Following expert review, duplicate, unclear, and clinically irrelevant items were removed, resulting in a 41-item trial version of the SBMAPRO comprising 27 physical function, 7 mental health, and 7 social functioning items.

### Stage 2: Item Selection and Development of the SBMAPRO

#### Participants

In Stage 2, 106 patients with SBMA completed the online survey and were included in the analysis.

#### Exploratory Factor Analysis and Internal Consistency

Exploratory factor analysis was conducted using maximum likelihood estimation with Promax rotation. Item selection was primarily based on factor loadings (≥0.40), while clinical interpretability was also considered when necessary (Online Resource 1). The resulting factor structure was largely consistent with the predefined conceptual domains.

Five latent factors were identified, corresponding to three physical function domains (bulbar symptoms, upper limb function, and trunk–lower limb function), one mental health domain, and one social functioning domain. Internal consistency, assessed using Cronbach’s alpha coefficients, demonstrated acceptable to excellent reliability across all domains, with values ranging from 0.651 to 0.901 (Table 1).

**Table 1.** Structural validity and internal consistency of the SBMA-PRO.

| | Number of items | Cronbach $\alpha$ |
| --- | --- | --- |
| Physical Function (bulbar symptoms) | 7 | 0.7959 |
| Physical Function (upper limb symptoms) | 4 | 0.7147 |
| Physical Function (trunk and lower limb symptoms) | 10 | 0.9055 |
| Mental Health | 6 | 0.6781 |
| Social Functioning | 4 | 0.6506 |

Exploratory factor analysis results and Cronbach's alpha coefficients for each domain of the SBMAPRO, demonstrating the multidimensional structure and internal consistency of the instrument.

#### Item Reduction and Scale Refinement

Based on the factor analysis results and evaluation of item performance, items were iteratively reviewed and reduced to improve clarity, relevance, and domain coherence.

As a result, the scale was refined to a total of 31 items, comprising 21 physical function items, 6 mental health items, and 4 social functioning items. In addition, response distribution patterns were reviewed, and to enhance scale usability and discrimination, the original 5-point Likert response format was revised to a 4-point scale.

#### Final Structure of the SBMAPRO

The final SBMAPRO demonstrated a three-subscale structure based on exploratory factor analysis. The physical function subscale included bulbar symptoms (7 items), upper limb symptoms (4 items), and trunk–lower limb symptoms (10 items). In addition, mental health (6 items) and social functioning (4 items) subscales were developed to comprehensively capture the multidimensional impact of SBMA. The complete SBMAPRO questionnaire, including all items and response options, is provided in Supplementary Table 2.

### Stage 3: Reliability and Validity Assessment of the SBMAPRO

The reliability and validity of the SBMAPRO were evaluated in two independent cohorts from Japan and the United States.

#### Japanese Cohort

##### Participants

Ninety Japanese patients with SBMA were included in the Stage 3 analysis. Demographic and clinical characteristics are summarized in Table 2.

**Table 2.** Demographic and clinical characteristics of the Japanese cohort (n = 90) and the U.S. cohort (n = 25)

|  | Japanese cohort (n=90) | U.S. cohort (n=25) |
| --- | --- | --- |
| | Average ± SD | Average ± SD |
| Age (years) | 58.5 ± 10.7 | 58.4 ± 10.1 |
| Age at onset (years) | 43.5 ± 10.4 | 41.2 ± 12.1 |
| Duration of disease (years) | 14.9 ± 7.6 | 17.2 ± 10.8 |
| Height (cm) | 168.7 ± 5.5 | 177.9 ± 6.2 |
| Weight (kg) | 63.5 ± 10.9 | 90.5 ± 18.7 |
| Number of CAG repeats | 47.9 ± 4.6 | 44.9 ± 3.1 |
| SBMAFRS | 39.7 ± 8.5 | 41.1 ± 7.1 |
| ALSFRS-R | 39.2 ± 5.2 | 40.2 ± 4.6 |

SD, standard deviation; SBMAFRS, spinal and bulbar muscular atrophy functional rating scale; ALSFRS-R, amyotrophic lateral sclerosis functional rating scale
Baseline demographic and clinical characteristics of Japanese and U.S. patients with spinal and bulbar muscular atrophy (SBMA) included in the Stage 3 analysis.

##### Reliability

Test–retest reliability of the SBMAPRO was assessed in the Japanese cohort. The ICCs demonstrated high reliability across all domains: 0.941 for physical functioning, 0.877 for mental health, and 0.858 for social functioning (Figure 2). The relationship between test and retest scores is shown in Figure 2.

**Fig. 2.**
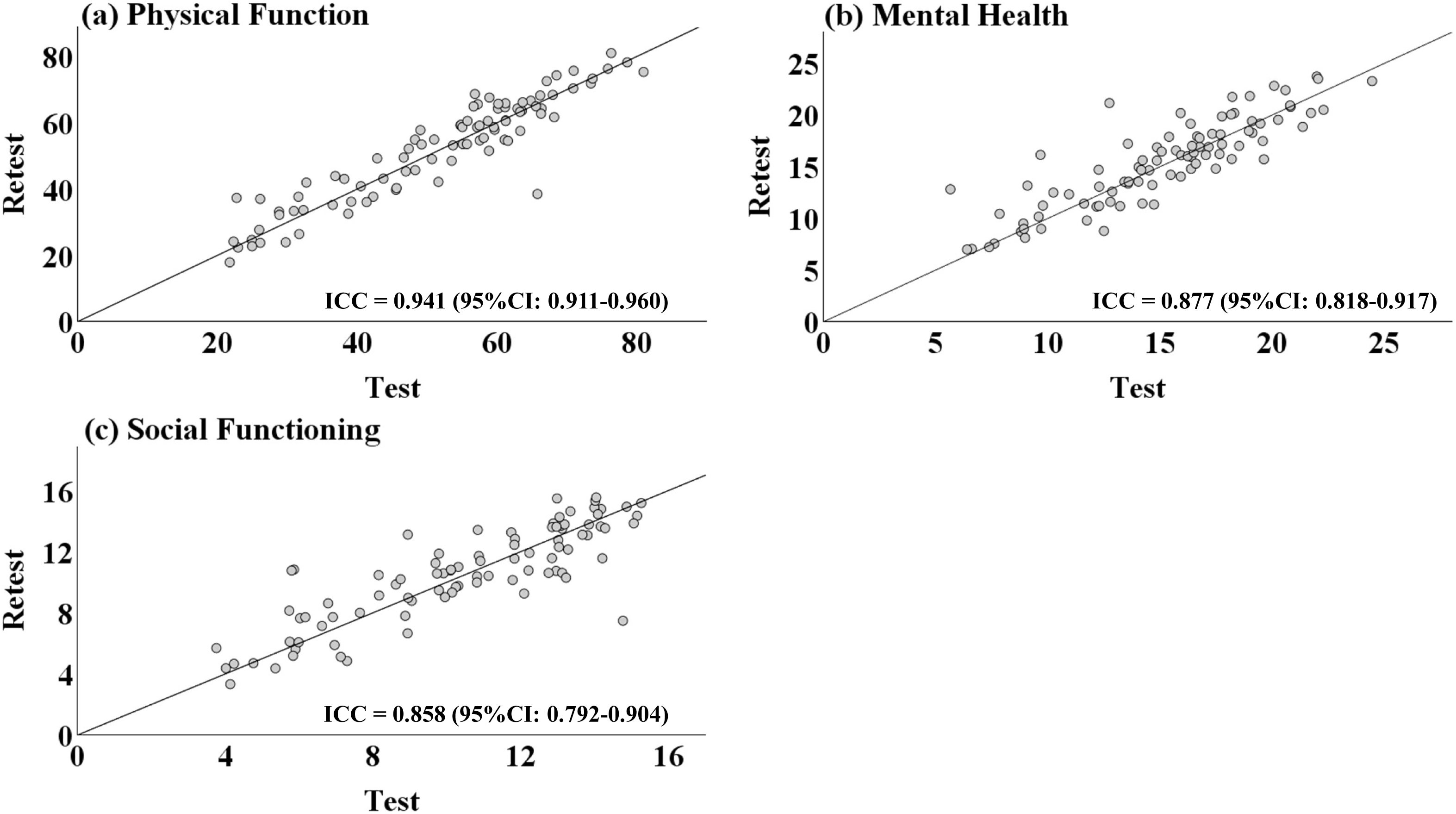
Test–retest agreement of SBMA-PRO domain scores in the Japanese cohort. Test–retest agreement plots for the SBMA-PRO domain scores in the Japanese cohort: (a) physical function, (b) mental health, and (c) social function

##### Validity

Construct validity was evaluated by examining correlations between SBMAPRO domain scores and established functional and HR-QoL measures. SBMAPRO domain scores showed strong correlations with the SBMAFRS (*r* = −0.826, *p* < 0.001) and the ALSFRS-R (*r* = −0.790, *p* < 0.001). Moderate correlations were observed with the SF-36 mental health (*r* = −0.603, *p* < 0.001) and social functioning (*r* = −0.617, *p* < 0.001) subscales (Figure 3).

**Fig. 3.**
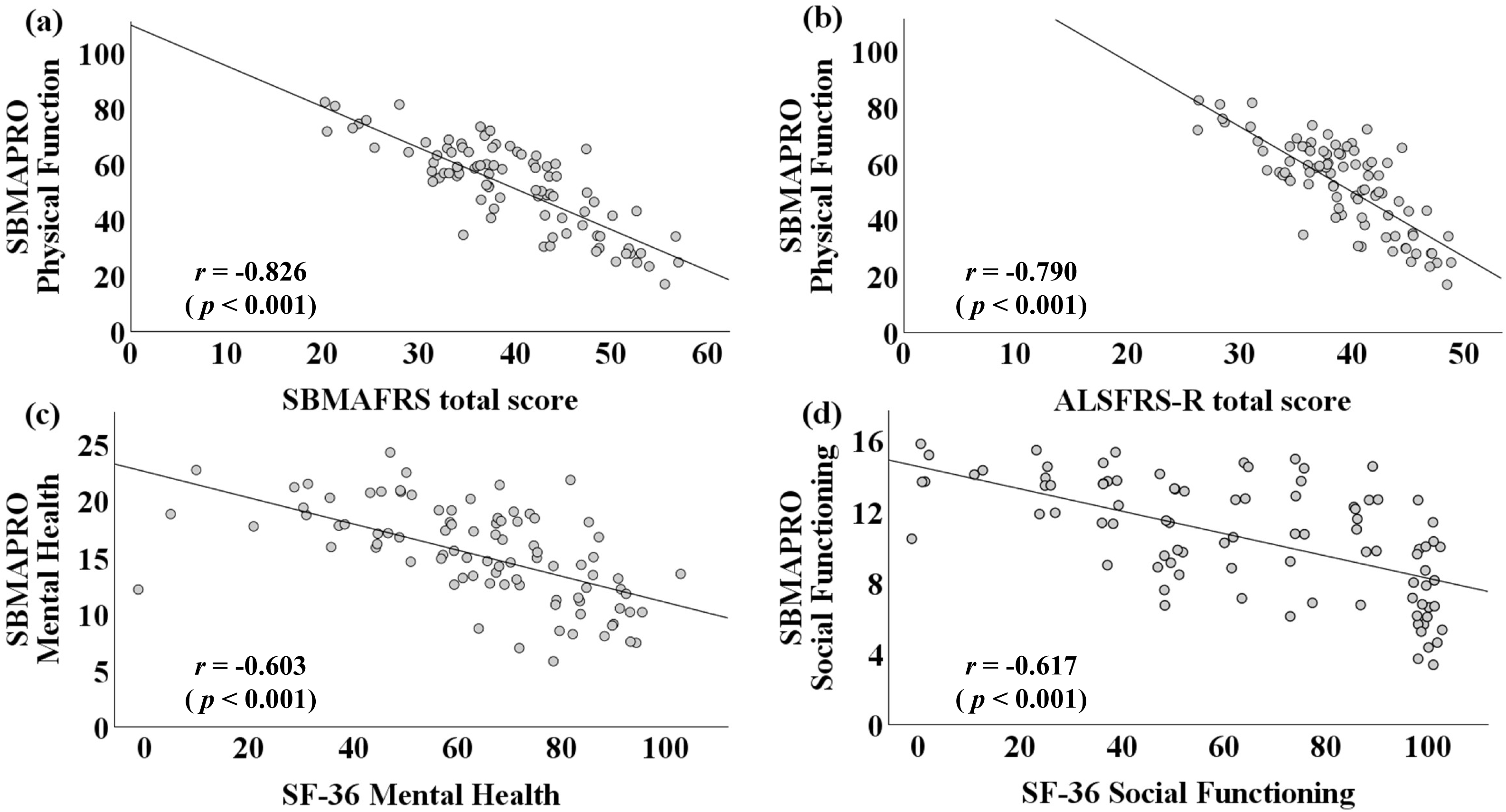
Construct validity of the SBMA-PRO in the Japanese cohort. Scatter plots illustrating correlations between SBMA-PRO domain scores and established functional and quality-of-life measures in the Japanese cohort: (a) SBMA Functional Rating Scale (SBMAFRS), (b) ALS Functional Rating Scale–Revised (ALSFRS-R), (c) SF-36 Mental Health, and (d) SF-36 Social Functioning

In subscale-level analyses, the mental health domain of the SBMAPRO demonstrated moderate correlations with the SBMAFRS total score (*r* = −0.416, *p* < 0.001) and the trunk–lower limb–related subscale score (*r* = −0.374, *p* < 0.001). The social functioning domain showed strong correlations with the SBMAFRS total score (*r* = −0.609, *p* < 0.001) and the trunk–lower limb–related subscale score (*r* = −0.587, *p* < 0.001). Complete correlations between SBMAPRO mental and social domains and SBMAFRS total and subscale scores in the Japanese cohort are presented in Table 3.

**Table 3.**
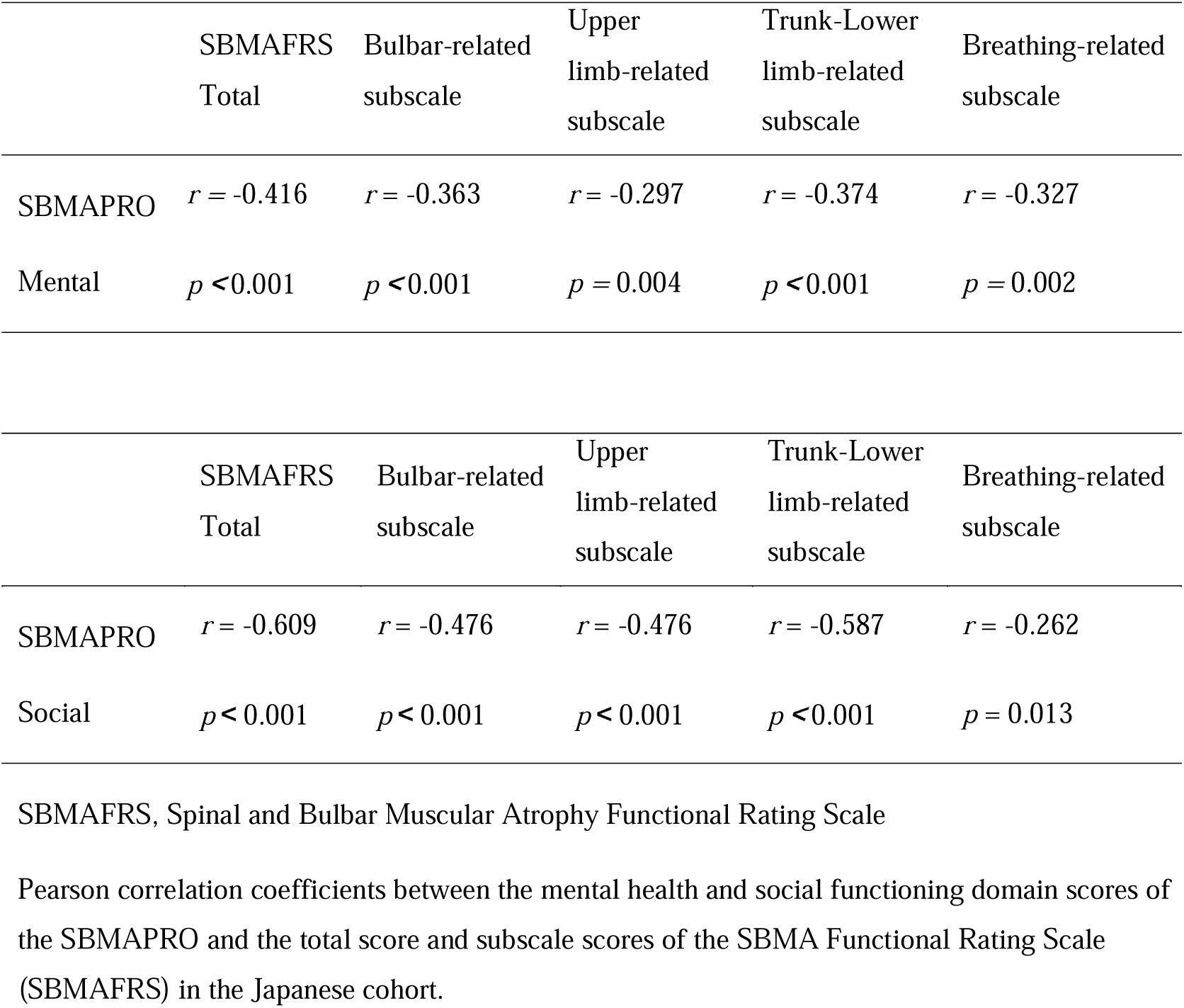
Correlations between SBMAPRO mental and social domains and SBMAFRS total and subscale scores in the Japanese cohort (n = 90)

#### U.S. Cohort

##### Participants

Twenty-five patients with SBMA were included in the U.S. cohort. Participant characteristics are summarized in Table 2.

##### Validity

In the U.S. cohort, SBMAPRO domain scores demonstrated a strong correlation with the SBMAFRS (*r* = −0.871, *p* < 0.001) and the ALSFRS-R (*r* = −0.647, *p* < 0.001). With respect to generic HR-QoL measures, the mental health domain of the SBMAPRO showed a strong correlation with the SF-36 mental health subscale (*r* = −0.829, *p* < 0.001), while the social functioning domain showed a moderate correlation with the SF-36 social functioning subscale (*r* = −0.627, *p* < 0.001) (Figure 4).

**Fig. 4.**
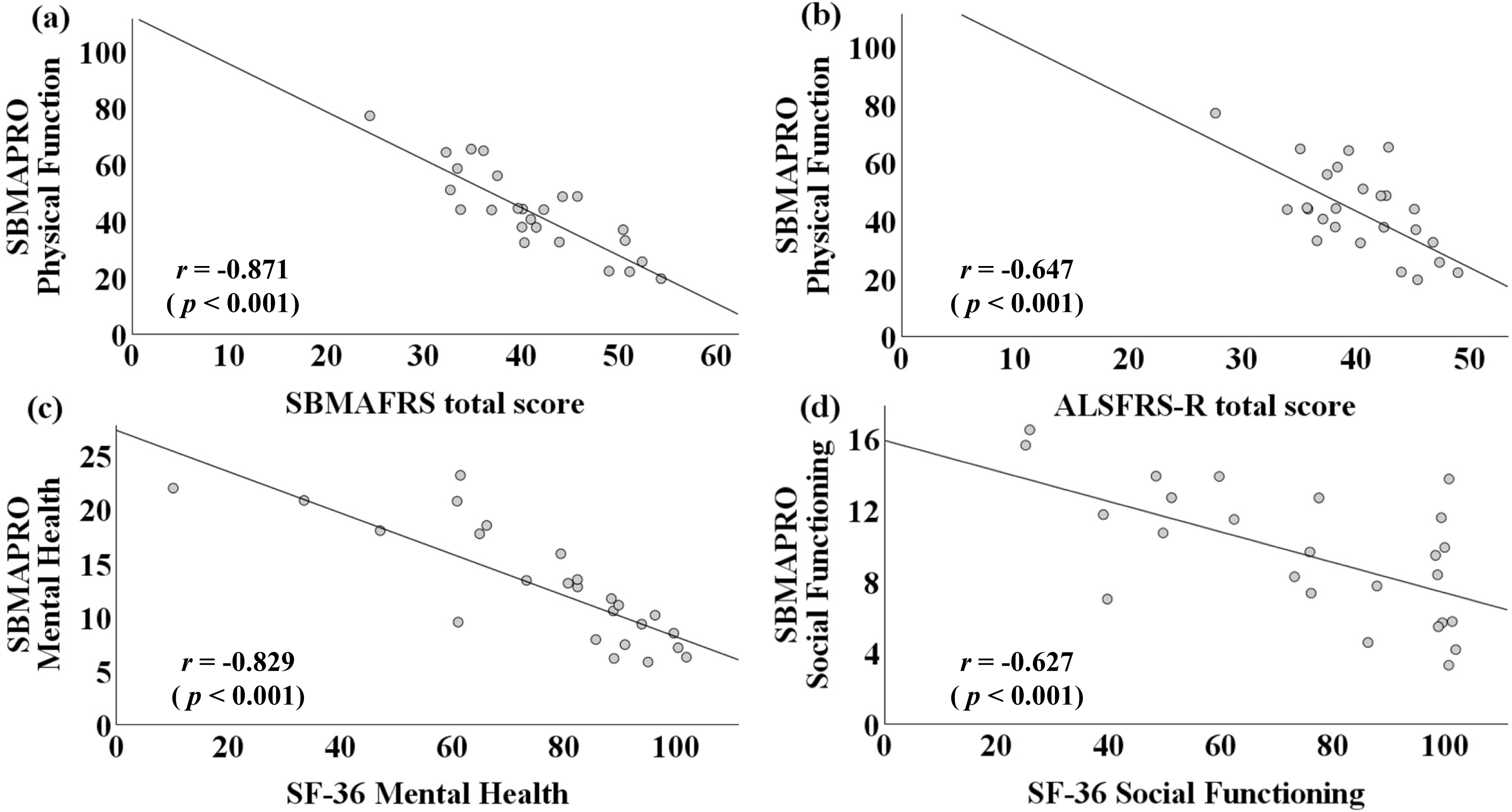
Construct validity of the SBMA-PRO in the U.S. cohort. Scatter plots illustrating correlations between SBMA-PRO domain scores and established functional and quality-of-life measures in the U.S. cohort: (a) SBMA Functional Rating Scale (SBMAFRS), (b) ALS Functional Rating Scale–Revised (ALSFRS-R), (c) SF-36 Mental Health, and (d) SF-36 Social Functioning.

Subscale-level analyses further demonstrated that, in the U.S. cohort, the mental health domain of the SBMAPRO was moderately correlated with the SBMAFRS trunk–lower limb–related subscale score (*r* = −0.515*, p* = 0.008), while the social functioning domain was strongly correlated with the same SBMAFRS subscale (*r* = −0.629, *p* < 0.001). Complete correlations between SBMAPRO mental and social domains and SBMAFRS total and subscale scores in the U.S. cohort are summarized in Table 4.

**Table 4.** Correlations between SBMAPRO mental and social domains and SBMAFRS total and subscale scores in the U.S. cohort (n = 25)

|  | SBMAFRS<br>Total | Bulbar-related<br>subscale | Upper<br>limb-related<br>subscale | Trunk-Lower<br>limb-related<br>subscale | Breathing-related<br>subscale |
| --- | --- | --- | --- | --- | --- |
| SBMAPRO | $r = -0.403$ | $r = -0.144$ | $r = -0.124$ | $r = -0.515$ | $r = -0.713$ |
| Mental | $p = 0.046$ | $p = 0.491$ | $p = 0.556$ | $p = 0.008$ | $p = 0.408$ |
|  | SBMAFRS<br>Total | Bulbar-related<br>subscale | Upper<br>limb-related<br>subscale | Trunk-Lower<br>limb-related<br>subscale | Breathing-related<br>subscale |
| SBMAPRO | $r = -0.538$ | $r = -0.301$ | $r = -0.082$ | $r = -0.629$ | $r = -0.327$ |
| Social | $p = 0.006$ | $p = 0.143$ | $p = 0.695$ | $p < 0.001$ | $p = 0.111$ |

SBMAFRS, Spinal and Bulbar Muscular Atrophy Functional Rating Scale
Pearson correlation coefficients between the mental health and social functioning domain scores of the SBMAPRO and the total and subscale scores of the SBMA Functional Rating Scale (SBMAFRS) in the U.S. cohort.

The strength of correlations was interpreted in accordance with commonly used guidelines in medical research [21].

## Discussion

The need for disease-specific PRO measures in SBMA arises from the distinct clinical features of this slowly progressive disorder. Although many patients maintain ambulation for decades, they experience early and persistent impairments in speech, swallowing, fatigue, emotional well-being, and social participation. These aspects may not be adequately reflected by clinician-reported functional scales alone. In the present study, we developed an SBMA-specific PRO in collaboration with patients. Our results demonstrate that the SBMAPRO captures important dimensions of the patient experience, complementing existing functional measures and addressing aspects of disease burden that have been difficult to quantify.

In this study, the SBMAPRO was developed and validated using a multi-stage framework designed to ensure content validity, structural validity, and reliability. Through qualitative patient interviews and expert review, a comprehensive item pool reflecting the physical, mental, and social impacts of SBMA was generated, establishing strong content validity. Subsequent exploratory factor analysis supported a multidimensional structure comprising physical function domains (bulbar, upper limb, and trunk–lower limb), along with mental health and social functioning domains. Internal consistency was satisfactory across all domains, supporting the reliability of the identified domain structure. Item performance was further evaluated during scale refinement to ensure coherence within each domain. The response format was further optimized by collapsing the original five-point scale into a four-category scale.

In the final validation phase, the SBMAPRO demonstrated high test–retest reliability in the Japanese cohort and showed good correlations with disease-specific functional measures in both the Japanese and U.S. cohorts, supporting construct validity. The relatively weaker correlations with generic QoL measures are consistent with the expectation that disease-specific instruments are more sensitive to SBMA-related impairments than generic tools. Notably, subscale analyses revealed consistent associations between mental health and social functioning scores and trunk–lower limb-related functional impairment, suggesting that limitations in mobility and postural control may substantially contribute to psychological burden and restricted social participation. These findings are consistent with the findings of a recent multicenter observational study showing that approximately 40% of patients reported walking and mobility as a primary problem [23].

A disease-specific PRO measure for SBMA, the SBMA-Health Index (SBMA-HI), has also recently been developed and validated [24]. Both the SBMA-HI and the SBMAPRO are comprehensive, disease-specific PRO instruments encompassing physical, mental, and social domains. However, whereas the SBMA-HI was primarily designed to quantify the overall burden of disease from the patient’s perspective, the SBMAPRO was designed with greater emphasis on a structured HR-QoL framework, enabling a more focused evaluation of relationships among physical, mental, and social domains.

The observed pattern of associations provides clinically meaningful insights into the multidimensional burden of SBMA. The present findings highlight the relevance of trunk and lower limb dysfunction to mental and social well-being beyond physical disability alone. These results underscore the importance of evaluating PRO measures to capture the impact of motor impairment on daily life, social roles, and autonomy, which may not be fully captured by clinician-rated scales. Differences in correlations with generic QoL measures observed between cohorts may reflect sample size limitations and cultural contexts.

From both clinical and research perspectives, the SBMAPRO represents a useful tool for supporting patient-centered care by incorporating the patient perspective into treatment planning and monitoring disease progression. It may also serve as an outcome measure in clinical trials aimed at detecting meaningful changes beyond clinician-reported function. The clear domain structure and simplified four-category response format enhance interpretability and may reduce respondent burden. Integration of the SBMAPRO into electronic data capture systems could further facilitate longitudinal assessment and patient engagement. The present study provides a foundation for future research focused on evaluating treatment responsiveness.

This study has several limitations. Item generation was based on participants from a single site, which may not fully capture regional variation. The psychometric analyses in Stage 2 were based on cross-sectional data, limiting the ability to assess responsiveness to change over time and to evaluate longitudinal associations between SBMAPRO and clinical measures. Consequently, the ability of the SBMAPRO to detect clinically meaningful changes or treatment effects remains to be determined, particularly for monitoring disease progression or evaluating therapeutic interventions. In addition, the U.S. cohort was limited in size, and test–retest reliability was not assessed. Future studies should focus on evaluating the responsiveness of the SBMAPRO through multisite longitudinal observational studies and on its application in clinical trials. These investigations should also examine clinically meaningful changes in SBMAPRO scores.

## Supporting information

Online Resource 1,2

## Data Availability

The datasets generated and/or analyzed during the current study are not publicly available.

## Acknowledgements

We sincerely thank all patients with SBMA and their families for their valuable participation in this study. We are grateful to the clinical staff at Nagoya University Hospital for their assistance with patient recruitment and data collection. We also acknowledge the support of the Neurogenetics Branch at the National Institute of Neurological Disorders and Stroke (NINDS), National Institutes of Health (NIH), in facilitating the U.S. cohort. The contributions of the NIH author(s) were made as part of their official duties as federal employees and are in compliance with agency policy requirements; accordingly, these contributions are considered works of the United States Government. However, the findings and conclusions presented in this paper are those of the author(s) and do not necessarily reflect the views of the NIH or the U.S. Department of Health and Human Services.

## Statements and Declarations

