## Supplementary material for "Development of patient-reported outcome for spinal and bulbar muscular atrophy": Online Resource 1,2

Online Resource **1. Exploratory Factor Analysis**

| **Oral functions** | 1 | 2 | 3 | 4 | 5 |
|---|---|---|---|---|---|
| FB 1. Have some trouble speaking | -0.077 | -0.098 | 0.689 | 0.153 | -0.002 |
| FB 2. Have nasal voice | -0.005 | -0.028 | 0.529 | -0.016 | 0.094 |
| FB 3. Can't chew hard foods | 0.044 | 0.024 | 0.759 | -0.168 | 0.166 |
| FB 4. Some pieces of food are left in the mouth after eating | -0.024 | -0.041 | 0.548 | 0.016 | 0.383 |
| FB 5. Choked when eating | 0.024 | 0.123 | 0.303 | -0.036 | 0.388 |
| FB 6. Slow eating speed | -0.036 | 0.209 | 0.652 | 0.064 | 0.086 |
| FB 7. Drooling even when not sleeping | -0.092 | -0.101 | 0.24 | -0.011 | 0.506 |

| **Upper limb functions** | 1 | 2 | 3 | 4 | 5 |
|---|---|---|---|---|---|
| FU 1. Can't lift heavy objects | 0.232 | -0.161 | 0.195 | 0.569 | 0.105 |
| FU 2. Can't use chopsticks, knife, and fork well | 0.177 | 0.072 | 0.045 | 0.238 | 0.444 |
| FU 3. Difficult to raise both arms | 0.265 | -0.247 | -0.004 | 0.333 | 0.556 |
| FU 4. Can't grip objects firmly | -0.005 | -0.068 | 0.004 | 0.708 | 0.295 |
| FU 5. Can't write well | -0.137 | 0.083 | 0.023 | 0.579 | 0.286 |
| FU 6. Difficult to button a shirt | 0.080 | 0.083 | 0.017 | 0.494 | 0.392 |
| FU 7. Have tremor in the hands | -0.329 | -0.003 | -0.111 | 0.392 | 0.276 |
| FU 8. Difficult to put strength into your hands when cold | -0.031 | 0.025 | 0.023 | 0.604 | -0.017 |
| FU 9. Can't turn a doorknob well | 0.152 | 0.135 | -0.054 | 0.212 | 0.425 |

| **Lower limb, trunk, and other functions** | 1 | 2 | 3 | 4 | 5 |
|---|---|---|---|---|---|
| FLT 1. Difficult to walk | 0.872 | 0.115 | 0.114 | -0.022 | -0.181 |
| FLT 2. Difficult to climb stairs | 0.834 | -0.203 | 0.089 | 0.239 | -0.329 |
| FLT 3. Difficult to go downstairs | 0.951 | -0.091 | -0.037 | -0.123 | 0.082 |
| FLT 4. Fall while walking | 0.805 | -0.003 | 0.016 | -0.245 | 0.135 |
| FLT 5. Have a knee-break when walking | 0.740 | 0.180 | -0.132 | -0.043 | 0.094 |
| FLT 6. Difficult to get up from a lying position | 0.497 | -0.016 | -0.107 | 0.211 | 0.203 |
| FLT 7. Difficult to stand up from a sitting position | 0.666 | 0.034 | 0.031 | 0.202 | -0.137 |
| FLT 8. Have numbness in extremities | -0.018 | 0.191 | 0.070 | 0.059 | 0.382 |
| FLT 9. Inconvenience when using the toilet | 0.675 | 0.101 | -0.065 | -0.165 | 0.32 |
| FLT 10. Inconvenience when taking a bath/ a shower | 0.481 | 0.135 | -0.046 | -0.044 | 0.374 |
| FLT 11. Leakage of stool or urine | 0.109 | -0.048 | 0.274 | -0.003 | 0.546 |

| **How you are feeling mood-wise about your disease** | 1 | 2 | 3 | 4 | 5 |
|---|---|---|---|---|---|
| M 1. Have no motivations to try new things | 0.013 | 0.685 | -0.013 | 0.029 | 0.106 |
| M 2. Pessimistic about the future | -0.154 | 0.883 | -0.026 | 0.101 | -0.08 |
| M 3. Increase irritability due to illness | -0.039 | 0.645 | -0.104 | 0.076 | 0.180 |
| M 4. Feel sorry for your family because your illness | 0.051 | 0.540 | 0.252 | -0.109 | -0.06 |
| M 5. Feel tired all the time | -0.041 | 0.547 | 0.280 | 0.078 | 0.015 |
| M 6. Feel difficult to tell the disease name to family members | -0.210 | 0.281 | 0.032 | -0.006 | -0.126 |
| M 7. Have decreased sexual desire and interest | 0.135 | 0.159 | 0.054 | 0.138 | 0.045 |

| The social impact illness may have on your life | 1 | 2 | 3 | 4 | 5 |
|---|---|---|---|---|---|
| S 1. Have less interaction with neighbors | 0.183 | 0.527 | -0.078 | 0.530 | -0.033 |
| S 2. Have experienced workplace harassment affecting work | 0.289 | 0.418 | -0.018 | -0.240 | -0.012 |
| S 3. I had to give up the work I wanted to do because of my illness | -0.112 | 0.425 | -0.043 | 0.480 | -0.026 |
| S 4. Income has decreased | 0.031 | 0.348 | 0.019 | 0.248 | -0.068 |
| S 5. Had to give up a hobby because my illness | 0.006 | 0.374 | -0.077 | 0.296 | -0.138 |
| S 6. My activity range has been reduced because my illness | 0.306 | 0.231 | -0.030 | 0.538 | -0.189 |
| S 7. Disagreement with my family because of my illness | 0.027 | 0.531 | -0.026 | -0.048 | 0.107 |

Online Resource **2. The SBMAPRO questionnaire (U.S. version)**

The complete SBMAPRO questionnaire, including all items organized by domain and the corresponding 4-point response scale.

| | **Completely disagree** | **Disagree/ Neither agree nor disagree** | **Slightly agree** | **Completely agree** |
|---|---|---|---|---|
| **I. Question about oral function** | | | | |
| **1-1. Have some trouble speaking** | O | O | O | O |
| **1-2. Have nasal voice** | O | O | O | O |
| **1-3. Can't chew hard foods** | O | O | O | O |
| **1-4. Some pieces of food are left in the mouth after eating** | O | O | O | O |
| **1-5. Choked when eating** | O | O | O | O |
| **1-6. Slow eating speed** | O | O | O | O |
| **1-7. Drooling even when not sleeping** | O | O | O | O |

| | Completely disagree | Disagree/Neither agree nor disagree | Slightly agree | Completely agree |
|---|---|---|---|---|
| II. Question about upper limb function | | | | |
| 2-1. Difficult to raise both arms | ○ | ○ | ○ | ○ |
| 2-2. Can’t write well | ○ | ○ | ○ | ○ |
| 2-3. Difficult to put strength into your hands when cold | ○ | ○ | ○ | ○ |
| 2-4. Can’t turn a doorknob well | ○ | ○ | ○ | ○ |

| | Completely disagree | Disagree/ Neither agree nor disagree | Slightly agree | Completely agree |
|---|---|---|---|---|
| **III. Question about lower limb, trunk, and other functions** | | | | |
| **3-1. Difficult to walk** | ○ | ○ | ○ | ○ |
| **3-2. Difficult to go downstairs** | ○ | ○ | ○ | ○ |
| **3-3. Have a knee-break when walking** | ○ | ○ | ○ | ○ |
| **3-4. Fall while walking** | ○ | ○ | ○ | ○ |
| **3-5. Difficult to get up from a lying position** | ○ | ○ | ○ | ○ |
| **3-6. Difficult to stand up from a sitting position** | ○ | ○ | ○ | ○ |
| **3-7. Have numbness in extremities** | ○ | ○ | ○ | ○ |
| **3-8. Inconvenience when using the toilet** | ○ | ○ | ○ | ○ |
| **3-9. Inconvenience when taking a bath/ a shower** | ○ | ○ | ○ | ○ |
| **3-10. Leakage of stool or urine** | ○ | ○ | ○ | ○ |

| | Completely disagree | Disagree/Neither agree nor disagree | Slightly agree | Completely agree |
|---|---|---|---|---|
| **IV. Question related to how you are feeling mood-wise about your disease** | | | | |
| **4-1. Pessimistic about the future** | O | O | O | O |
| **4-2. Increase irritability due to illness** | O | O | O | O |
| **4-3. Feel sorry for your family because your illness** | O | O | O | O |
| **4-4. Feel tired all the time** | O | O | O | O |
| **4-5. Feel difficult to tell the disease name to family members** | O | O | O | O |
| **4-6. Have decreased sexual desire and interest** | O | O | O | O |

| | Completely disagree | Disagree/ Neither agree nor disagree | Slightly agree | Completely agree |
|---|---|---|---|---|
| **IV. Question regarding the social impact the illness may have on your life** | | | | |
| **5-1. Had to give up the work I wanted to do because of my illness** | ○ | ○ | ○ | ○ |
| **5-2. Had to give up a hobby because my illness** | ○ | ○ | ○ | ○ |
| **5-3. My activity range has been reduced because my illness** | ○ | ○ | ○ | ○ |
| **5-4. Disagreement with my family because of my illness** | ○ | ○ | ○ | ○ |
